# A quantitative tool for seizure severity: diagnostic and therapeutic applications

**DOI:** 10.1101/2022.10.26.22281569

**Authors:** Akash R. Pattnaik, Nina J. Ghosn, Ian Z. Ong, Andrew Y. Revell, William K.S. Ojemann, Brittany H. Scheid, John M. Bernabei, Erin Conrad, Saurabh R. Sinha, Kathryn A. Davis, Nishant Sinha, Brian Litt

## Abstract

**Objective:** More than one-third of the people with focal epilepsy do not achieve seizure freedom with medication, neuromodulation, or neurosurgery therapies. Palliative care with the goal of reducing epilepsy burden is an alternative for these patients. Minimizing severe seizures is essential for reducing morbidity. Existing seizure severity scales are qualitative and rely on patient reports, limiting our ability to rigorously track and intervene to curb severe seizures. The goal of this study is to develop and validate a quantitative metric for seizure severity.

**Methods:** We retrospectively analyzed preictal and ictal intracranial-EEG (iEEG) recordings from 54 people with drug-resistant epilepsy undergoing pre-surgical evaluation. We developed a new metric that objectively combines seizure duration, spread, and semiology to quantify seizure severity. We calculated preictal iEEG network features and fit a linear mixed-effects model to quantify patient-specific associations between preictal networks and seizure severity.

**Results:** We evaluated 256 seizures from 54 patients using the quantitative seizure severity score. Seizure severity was consistent with clinical seizure type. Medication taper strategy was associated with seizure severity (p = 0.018, 97.5% confidence interval = [-1.242, -0.116]) and lower pre-surgical seizure severity was associated with better post-surgical seizure outcome (U = 465, *p* = 0.042). A linear mixed-effects model with preictal network features as regressors and seizure severity as response revealed a group-level positive trend. In 12 out of 14 patients with multiple types of seizures, more severe seizures were preceded by more abnormal preictal networks.

**Significance:** We present a quantitative metric for seizure severity that correlates with clinical and electrographic features. We found that the seizure severity score was associated with abnormal preictal networks. We propose this measure to holistically capture patient condition and guide incremental changes in therapy to improve patient outcome over time.

## Introduction

Of the approximately 70 million people with epilepsy worldwide, more than 30% are resistant to anti-seizure medications (ASMs)^1^. For many of these patients, clinicians aim for palliative rather than curative therapies. These approaches focus on reducing the frequency and severity of seizures as clinicians adjust ASMs or expand ablation, resection, and stimulation targets to deliver better seizure control. There are currently no good measures of seizure severity to guide therapy changes other than patient seizure diaries, which can be inaccurate for many individuals^2^. A quantitative measure of seizure severity is essential for ensuring that therapies are effectively treating individuals with epilepsy.

Assessment of the clinical response to treatment relies mainly on seizure diaries in which patients report seizure type, semiology, duration, and frequency. Emerging evidence suggests that individuals with impaired awareness during their seizures may not be able to report epileptic events or their severity accurately^3–5^. A combination of at least three factors can characterize seizure severity: a) seizure duration, b) patterns of seizure spread, and c) seizure semiology. Seizure semiotics such as convulsions, lack of awareness, falls, disruptive automatisms, or seizure-related injuries are associated with decreased quality of life and increased risk of sudden unexpected death (SUDEP)^5–8^. Previous studies have quantified seizure severity from clinical semiology using survey-based scales^8–12^. The National Hospital Seizure Severity Scale (NHS3) is one such scale, amongst others, that is useful for measuring the efficacy of ASMs in clinical trials^8,12^. Though these scales quantify clinical semiology to estimate seizure severity, survey-based scales are limited by interpretation bias and insufficiently capture objective measures of severe seizures^12^.

Electrophysiological features characterizing seizure severity, such as seizure spread patterns and seizure duration, can be quantified from EEG. In epilepsy monitoring units (EMUs), patients are monitored with intracranial EEG (iEEG) for days to weeks, and seizures are induced by progressively withdrawing ASMs, sleep deprivation, or other provoking measures. Even in the controlled EMU setting, severe seizures carry a significant risk of injury^13,14^, so it is critical to minimize seizure severity while maximizing the diagnostic yield of iEEG. There is some precedent for using EEG measures of seizure severity to guide treatment; for example, in generalized epilepsies, long runs of primary generalized discharges, a measure of seizure duration, are associated with impaired awareness^15^. Developing a quantitative seizure assessment tool that combines clinical and electrophysiological features of seizures would be instrumental in guiding seizure reduction therapies.

In this study, we propose a quantitative measure of seizure severity that integrates seizure duration, spread, and semiology. We apply this seizure severity score to more than 250 seizures from 54 patients who underwent iEEG monitoring, many of whom exhibited multiple seizure types. We validate the sensitivity of the seizure severity score against clinical variables such as seizures after medication tapering, duration of epilepsy, seizure classification, and surgical outcomes. We finally test the hypothesis that quantitative measures derived from preictal iEEG recordings may predict more severe seizures and could notify patients and clinicians to preempt seizure-related injuries in the EMU.

## Materials and methods

### Patient characteristics

We retrospectively analyzed data from 256 seizures recorded in 54 individuals with epilepsy from the Hospital of the University of Pennsylvania (HUP). Data collection for research received prior approval by the HUP institutional review board and informed consent was obtained from each subject. Subjects completed (1) in-patient iEEG monitoring in the EMU, (2) pre- and post-implant MR and CT imaging, respectively, to localize electrode contacts, (3) resection or ablation surgery for seizure control, and (4) assessment of surgical outcome at one year after surgery. Patient demographics are provided in Table 1 and Table S1, dichotomized to determine clinical factors that are associated with FBTCS. All seizures were classified using the ILAE classification as focal aware seizures (FAS, *n* = 50), focal impaired awareness seizures (FIAS, *n* = 93), and focal to bilateral tonic-clonic seizures (FBTCS, *n* = 73)^16^. Subclinical seizures (SCS, *n* = 36) were identified on the EEG by the attending epileptologists and were accompanied by no obvious clinical symptoms. Four seizures were marked as unknown classification as ictal testing for awareness was not performed.

**Table 1.** Subject demographics and clinical data. Clinical variables for the subjects included in the study are given. Fisher’s exact test with *p* values for significance are reported.

|  | **FBTCS -** | **FBTCS +** | **Fisher's exact test (*p*)** |
| --- | --- | --- | --- |
| **Therapy**<br>( Resection / Ablation ) | ( 8 / 12 ) | ( 20 / 14 ) | 2.14 (0.26) |
| **Implant**<br>( ECoG / SEEG ) | ( 5 / 15 ) | ( 18 / 16 ) | 0.30 (0.05) |
| **Laterality**<br>( L / R ) | ( 15 / 5 ) | ( 18 / 16 ) | 2.67 (0.15) |
| **Lesion Status**<br>( Lesional / Non-lesional ) | ( 8 / 12 ) | ( 17 / 17 ) | 0.67 (0.58) |
| **Gender**<br>( F / M ) | ( 11 / 9 ) | ( 14 / 20 ) | 1.74 (0.40) |
| **Total** | **20** | **34** |  |

### iEEG data selection and processing

iEEG data were collected at 500, 512, or 1024 Hz using electrocorticography (ECoG) grids and strips, stereoelectroencephalography (SEEG) depth electrodes, or a combination of both modalities. In each patient, we segmented iEEG recordings into three categories: 1) ictal epochs for each seizure event, 2) 60-sec preictal epochs preceding each seizure’s onset, and 3) 20 interictal epochs, each 30-sec in duration, to quantify a baseline distribution.

Ictal epochs were identified from the clinical notes of the attending EMU epileptologist and confirmed with video-EEG. Interictal epochs were selected randomly and fulfilled the following criteria: clips were recorded 1) during awake segments of iEEG recording for consistency across patients, 2) more than 72 hours after implant to minimize implant effect, 3) at least 2 hours before annotated seizures, and 4) at least 2, 6, and 12 hours after subclinical, focal onset, and focal-to-bilateral seizures, respectively. Wake status was determined by a validated sleep detector^17^. In all recordings, we excluded the contacts in white matter, outside the brain, and corrupted with noise. On each channel, we applied a 60 Hz second-order IIR notch filter (quality factor 30) to remove power line noise and a third-order Butterworth filter between 1 to 120Hz to detrend and remove high-frequency noise, both in forward and backward directions to prevent artificially introduced phase shift. We applied a common-average reference to all channels.

On the preictal and interictal segments, we computed iEEG functional connectivity networks. We down-sampled the preictal and interictal epochs to 200Hz to maintain a consistent sampling rate across patients and applied a third-order Butterworth filter between 0.5 to 80Hz. We computed broadband coherence between each pair of electrodes using Welch’s method and a 2-s Hamming window with 1-s overlap. Channels were represented as nodes and broadband coherence between pairs of channels were represented as edges. We finally summarized the network topology in each epoch by computing the mean node strength across all channels.

To estimate the deviations in the preictal networks from the interictal baseline in each patient, we computed the mean and standard deviation of node strength from 20 interictal epochs. We *z*-scored preictal node strength using the interictal node strength mean and standard deviation, then computed the absolute value of the z-score. A higher (lower) preictal *z*-score meant that the preictal network was less (more) similar to the interictal baseline distribution.

### Seizure severity score

We obtained a seizure severity score for each seizure by combining the seizure duration, seizure spread pattern, and seizure semiology as shown in Figure 1 and detailed below:

**Figure 1.**
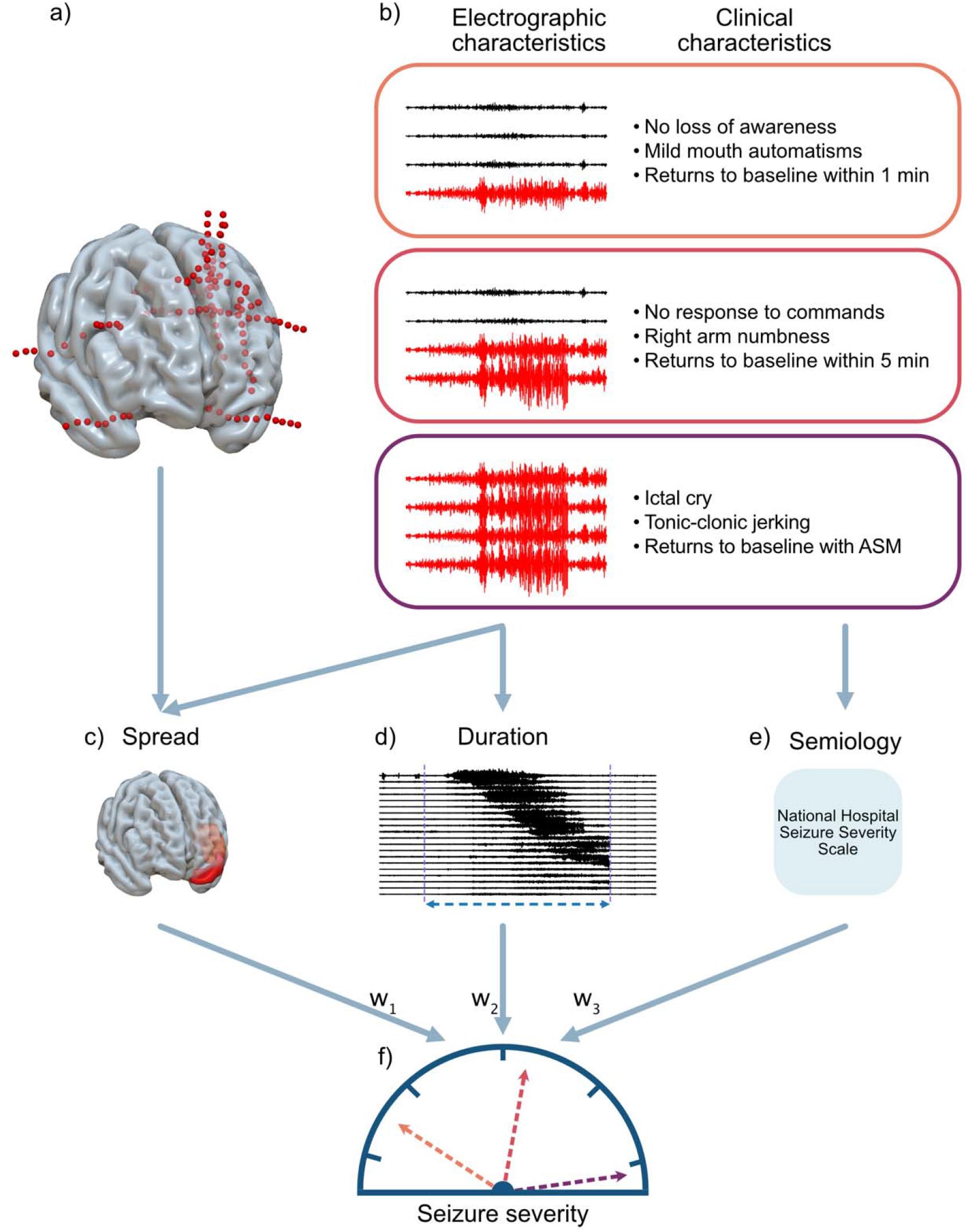
Seizure severity is composed of metrics for seizure spread, duration and semiology. (a) Patients are implanted with intracranial EEG to localize seizure onset. (b) Multiple seizures are recorded electrographically and corresponding clinical notes document semiology. (c) We quantify seizure spread as the number of brain regions to which a seizure has spread, (d) seizure duration as the time elapsed from earliest electrographic change to offset and (e) seizure semiology using a previously validated clinical scale. (f) Seizure spread, duration and semiology are objectively combined into one metric that represents seizure severity.

### Quantifying seizure duration and spread from electrographic characteristics

We quantified *seizure duration* from the clinical annotations of the iEEG recording. A clinical team of board-certified epileptologists visually reviewed iEEG data and marked the earliest electrographic change (EEC) and offset^18^. We computed seizure duration as the number of seconds from EEC to offset and transformed the duration to a log scale.

We quantified *seizure spread* by first separating iEEG contacts as those involved and not involved in the seizure using an absolute slope-based seizure detector. The absolute slope is a time-series metric that records both high-amplitude slow-wave and low-amplitude fast-wave signal properties, which are typical in iEEG recordings of seizures^19^ (see Supporting Information). Figure S1 shows that results obtained using the absolute slope method are consistent with results obtained using deep-learning-based seizure spread algorithms^20^. Next, we localized iEEG contacts to brain regions using a predefined Desikan-Killiany atlas^21^ by applying our previously published semi-automatic pipeline^22,23^. This pipeline registers post-implant CT imaging to pre-implant MRI, labels contacts in native space, and transforms the electrode coordinates in ICBM152 2009c non-linear symmetric standard space^24,25^. We constructed a 5mm sphere around each contact^26^ and mapped the contact to the overlapping brain regions defined by the atlas. The number of regions that overlapped with at least one seizing channel quantified the extent of seizure spread for every seizure.

### Quantifying seizure semiology from clinical characteristics

We quantified *seizure semiology* from the clinical features of each seizure by applying the previously validated NHS3 seizure-severity scale^8,12^. For each seizure, we extracted the clinical semiology and patient descriptions reported by epileptologists in their notes. Three independent reviewers rated each seizure on the 27-point NHS3 scale of seizure severity, which consists of questions related to fractures, falls, and automatisms, among other physical and social manifestations of seizures. The methods section in the Supporting Information shows the score sheet of the NHS3 seizure-severity scale.

### Integrating electrographic and clinical characteristics

Seizure duration, spread, and semiology were integrated into a single severity score for each seizure. We reduced the three features into a single seizure severity score using non-negative matrix factorization (NMF) for dimensionality reduction. NMF quantified the goodness-of-fit (R^2^) to interpret each feature’s contribution to the seizure severity score (see Supporting Information for details on the application of NMF).

## Statistical methods

To validate the seizure severity score and assess its sensitivity in response to changes in clinical therapy and disease severity, we tested its association with a) seizure type, b) epilepsy duration, c) ASM tapering load in the EMU, and d) seizure outcomes at one-year after surgery. Across all analyses, we chose nonparametric statistical tests over parametric methods to avoid making any assumptions about the distribution of features across seizures or patients. We hypothesized that more severe seizures would be associated with severe clinical seizure classification, longer epilepsy duration, lower ASM load due to medication tapering, and poor seizure outcomes. We rejected the null hypotheses for p < 0.05 and reported the test statistic for each analysis.

To test if seizure severity scores were different across seizure types, we applied the Kruskal-Wallis test and the post-hoc Dunn’s test for pairwise comparisons. One-tailed Spearman rank correlation was used to test the associations between epilepsy duration and patient-level severity, defined as the maximum seizure severity score across a patient’s seizures. We converted the ASM load in patients for each day in the EMU to a continuous metric by applying a first-order pharmacokinetics model, thereby quantifying the average ASM load for up to one hour before seizure onset. A linear mixed-effects model with ASM load as the fixed effect predictor, patient number as the random effect, and severity as the response was used to determine the effect of ASM load on severity. A Mann-Whitney rank sum test was used to determine if the distribution of patient-level severity was significantly different between subjects who were and were not seizure-free after surgery.

To test whether iEEG node strength in the preictal segments could predict seizure severity scores, we applied a linear mixed-effects model with random slope and intercept. We set the mean preictal node strength z-scores as the fixed effect predictor, individual patients as the random effect, and severity as the response to determine the patient-level associations between preictal deviation in node strength and seizure severity.

### Data availability

To ensure transparency and allow others to use our methods, we have made all our code available at https://github.com/penn-cnt/Pattnaik-seizure-severity. All iEEG recordings are accessible from iEEG.org.

## Results

### Seizure severity scores highlight the variability of seizures within patients

Across 256 seizures from 54 patients, seizure severity scores ranged from 0.03 to 2.3, with a median of 0.74 (Figure 2a). Seizure severity scores were bounded such that the lowest possible score was 0. We calculated the range of severity scores for each patient and found that severity scores varied substantially even in the same patient (median = 0.33, IQR = 0.79, Figure 2b).

**Figure 2.**
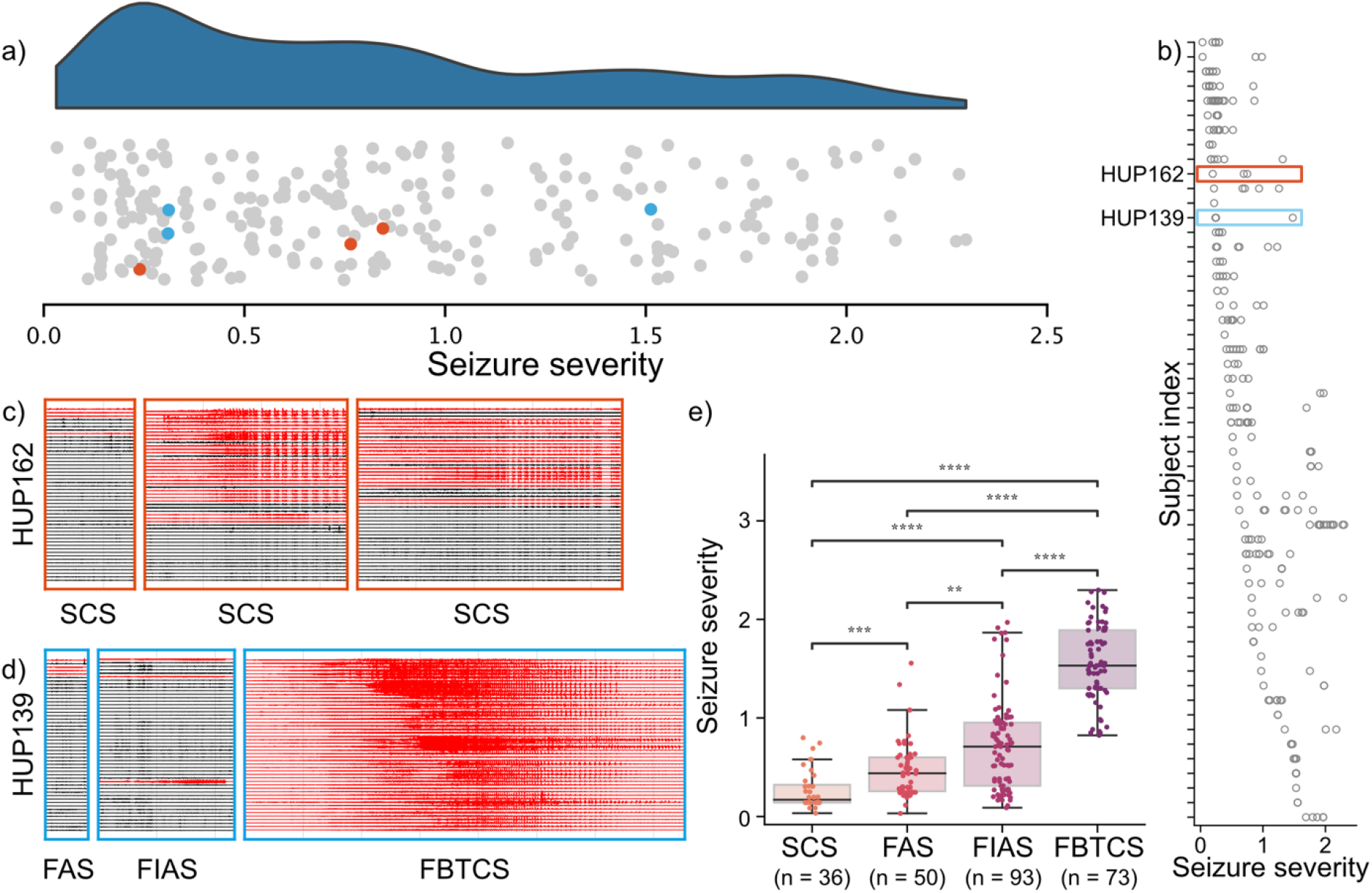
The seizure severity score quantifies differences within and across patients. (a) The rain cloud plot shows the distribution of severity values. Each point represents one seizure and example patients are highlighted in red and blue, respectively. The blue cloud shows a kernel density estimate. (b) Patient specific severity values. Patients are sorted by minimum severity value and example patients are highlighted in red and blue, respectively. (c) Seizure recordings of HUP162 with three labeled sub-clinical seizures (SCS). (d) Seizure recordings of HUP139 with a focal aware seizure (FAS), focal impaired aware seizure (FIAS), and a focal-to-bilateral-tonic-clonic seizure (FBTCS). Channels in red were detected as being part of the seizure and channels in black were detected as being spared from the seizure. (e) A plot of seizure severity by seizure type across all patients. (Kruskal-Wallis test with post-hoc Dunn’s test, *p* <0.05 for all pairs).

We highlight two example cases, HUP162 (in red) and HUP139 (in blue). All three seizures for HUP162 were deemed SCS, whereas HUP139 had an episode of FAS, FIAS, and FBTCS each. All seizures were deemed as subclinical for HUP162, though seizure spread patterns and duration were different (Figure 2c). This indicated different seizure severity (seizure 1 = 0.20, seizure 2 = 0.69, seizure 3 = 0.75). For HUP139, the difference in seizure semiology is apparent for seizure traces of the FBTCS characterized by longer duration and spread (Figure 2d). However, we found that the FAS and FIAS were more similar when seizure spread, duration, and other NHS3 clinical characteristics were factored in, resulting in seizure severity scores of 0.25 and 0.24 for the FAS and FIAS, respectively, and 0.75 for the FBTCS.

Figure 2e shows the seizure severity score for all seizures categorized by clinical seizure type. We highlight three observations from this plot: a) median seizure severity scores monotonically increase from SCS, FAS, FIAS, and FBTCS types, b) severity values vary even within a single seizure type, and c) distributions of severity overlapped across seizure types i.e., some seizures deemed as SCS, FAS, and FIAS, typically considered less severe, had higher scores than some FBTCS seizures, typically considered more severe. We detected a similar trend in Figure S2 upon categorizing individual features constituting seizure severity by seizure type.

Overall, these results suggest that our proposed seizure severity score is more sensitive in detecting variability between seizures, even in the same patient, than the gross classification of seizures by clinical semiology alone.

### Seizure severity score is sensitive to changes in disease severity and clinical response

Severe seizures are a hallmark of severe epileptic disorders^27^; therefore, we tested if our proposed seizure severity score correlates with factors known to be associated with epilepsy severity. Specifically, we focused on three independent factors: a) *epilepsy duration*: patients who experience uncontrolled seizures for longer may exhibit greater changes in their epileptic networks and more severe seizures^22,28^, b) *ASM load taper*: more severe seizures might occur when patients are weaned off ASMs in the EMU^29^, and c) *seizure outcomes*: poor seizure outcomes (Engel class 2-4) after surgery are associated with widespread abnormalities in epileptic network and may permit severe seizures before surgery^30^.

Epilepsy severity, an important indicator of cognitive and social burden, consists of seizure frequency, seizure severity, and the degree to which brain networks are altered. Figure 3 shows the association of these three independent clinical factors with each patient’s seizure severity score. We found that seizure severity correlated with epilepsy duration (Spearman *r* = 0.23, *p* = 0.049), ASM load taper (p = 0.018, 97.5% confidence interval = [-1.242, -0.116]), and seizure outcomes (U = 465, *p* = 0.042). These results appear to validate our hypotheses that more severe seizures might occur with longer epilepsy duration and at lower doses of ASMs in the EMU. The association of more severe seizures with one-year seizure outcomes after surgery suggests that the seizure severity score might be a sensitive predictor of response to therapy.

**Figure 3.**
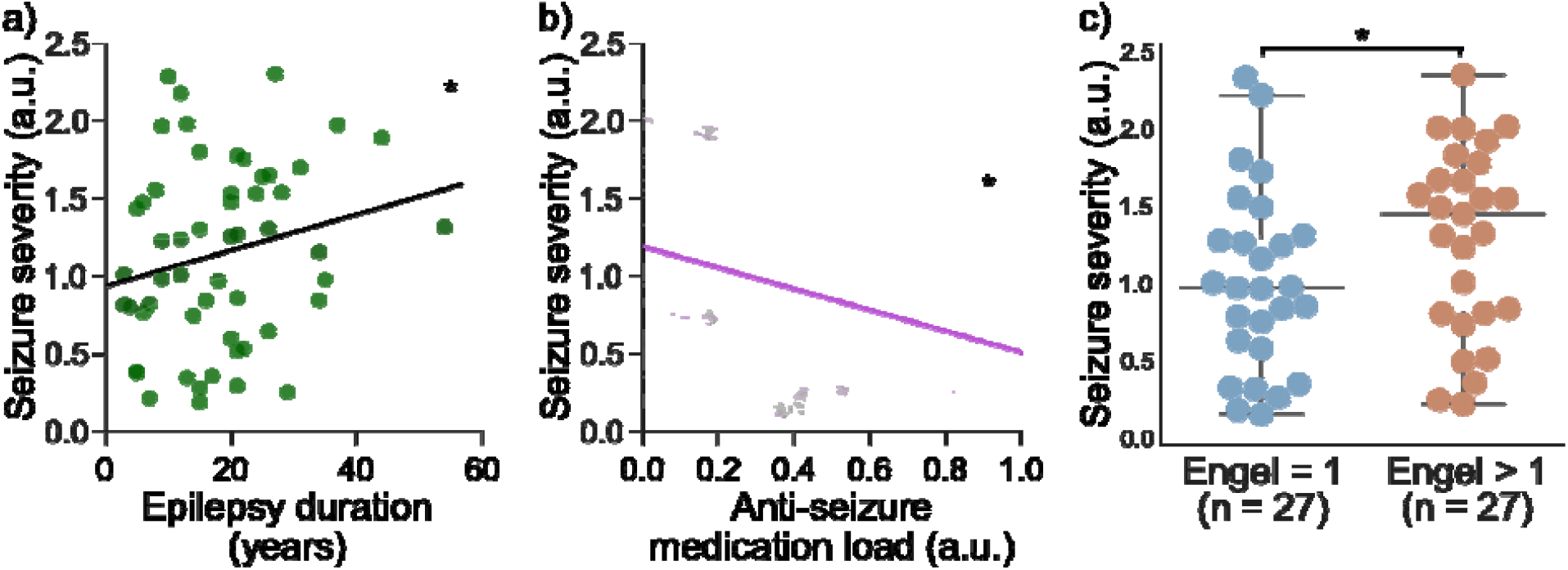
Seizure severity is associated with clinical variables of epilepsy. (a) Epilepsy duration is correlated with maximum seizure severity (one-tailed Spearman’s *r* = 0.22, *p* = 0.05). Individual points represent patients. Linear regression line and 95% confidence interval are shown in black and gray. (b) A linear mixed effects model was fit with anti-seizure medication as the covariate, patient as the random effect, and seizure severity score as the response variable (p = 0.018, 97.5% confidence interval = [-1.242, -0.116]). The thick purple line shows the fixed effects intercept and slope, and the thin purple lines show each patient’s random intercept. Individual data points represent each seizure and are plotted in grey. (c) Poor outcome patients are associated with more severe seizures than good outcome patients. The most severe seizure within each patient was used to measure patient-level severity (Mann-Whitney *U* = 264, *p* = 0.04).

### Deviations in preictal iEEG network node strength from baseline are associated with seizure severity within patients

We assessed if preictal iEEG features could predict the severity of impending seizures in individual patients. Premising on emerging evidence that distinct changes in iEEG functional connectivity precede seizures of different severity^31^, we identified patients whose seizure severity scores had a variance of at least 0.1 (*n* = 14). These patients had seizures in the EMU that were very dissimilar, spanning a wide range on the spectrum of the seizure severity score.

We computed the deviation of iEEG functional connectivity node strength in the preictal segment preceding each seizure from the seizure-free/interictal baseline. Modeling the relationship between seizure severity score and node strength, we found that most patients had a positive trend. At the group level, there was a significant positive association between preictal deviation in node strength and seizure severity (beta = 0.68, 95% CI = [0.12, 1.25]).

The positive association, *at the group level*, demonstrates that preictal iEEG segments with greater deviations in node strength precede more severe seizures in approximately 86% (12/14) of patients. However, the opposite effect between seizure severity and preictal deviation in node strength *at the individual patient level* (Figure 4b-c) suggests that network changes prior to seizure onset might not fully explain the occurrence of severe seizures in 14% (2/14) of our cohort. Analyzing chronic iEEG recordings and a larger sample size are needed to better understand the influence of preictal dynamics on seizure severity at a patient-specific scale.

**Figure 4.**
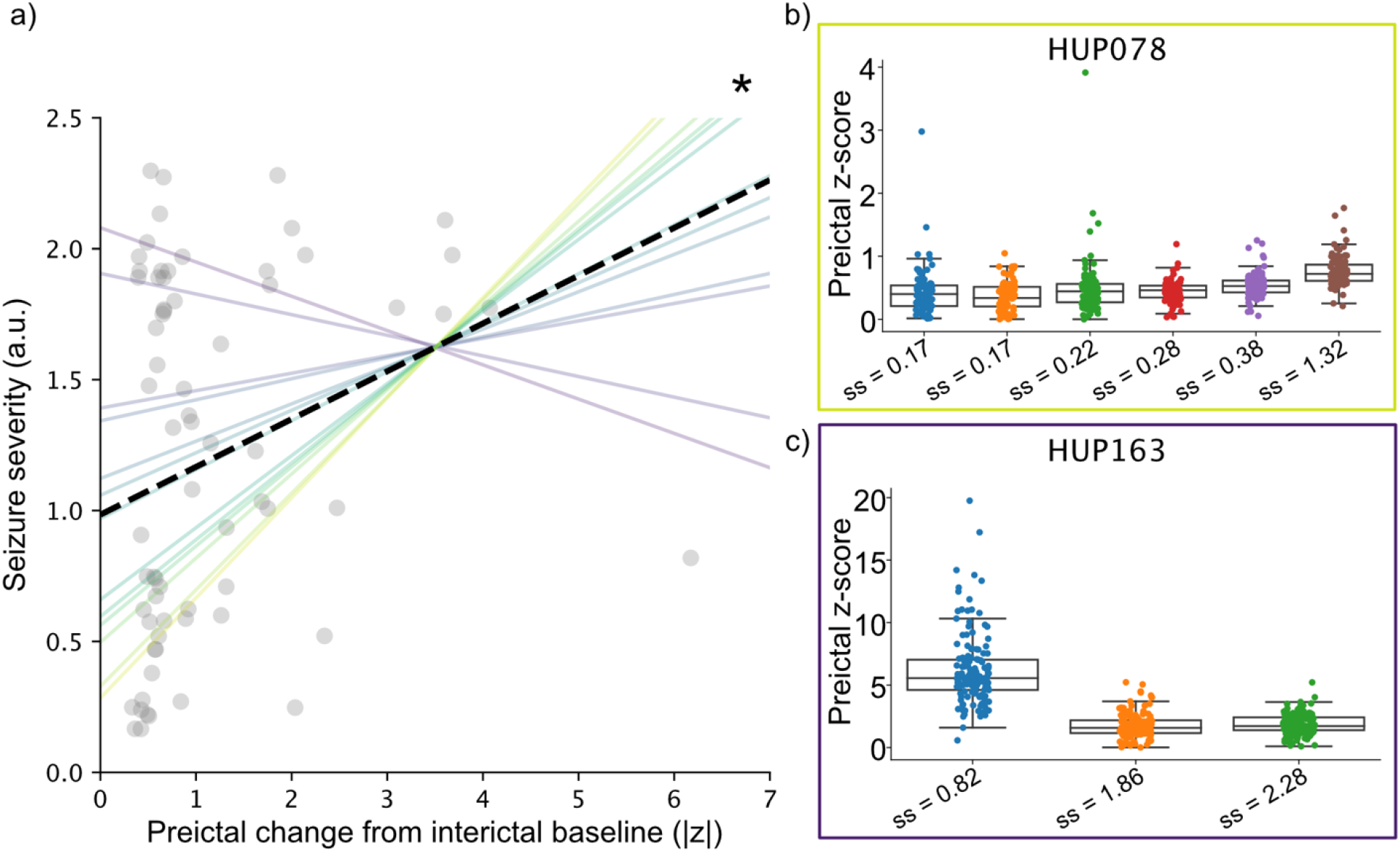
Patient-level trends in preictal node strength and severity. (a) A linear mixed effects model was fit with preictal bivariate features as the covariate, patient as the random effect, and seizure severity score as the response variable (p = 0.034, 97.5% confidence interval = [0.013, 0.352]). The thick dashed line shows the fixed effects intercept and slope, and the thin purple lines show each patient’s random intercept and slope. Individual data points represent each seizure and are plotted in grey. (b) Electrode-level features from the patient with the largest positive slope. Boxplots summarize electrode-level features and seizures are sorted by increasing severity. (c) Same layout as (b) for the patient with the largest negative slope.

## Discussion

Quantifying seizure severity is critical to assess disease burden in epilepsy and measuring response to therapy, particularly for neurostimulation and medications which are adjusted over multiple years. We propose a new metric to quantify seizure severity that builds on the previous semiology-based seizure severity scales by including electrophysiological features that characterize seizure spread and duration. The proposed seizure severity score highlights seizure-specific variability in individual patients and demonstrates the overlap between clinical seizure classifications. By assessing the association between seizure severity scores and independent clinical measures related to epilepsy severity, we found that our seizure severity score might be sensitive to changes in clinical response. We investigated the iEEG data before seizures to suggest network measures that may predict the likelihood of severe seizures that could help preempt seizure-related injuries.

Seizure frequency and type are typically recorded from patient diaries maintained by people with epilepsy, and have been shown to be inaccurate and inconsistent for children and people who have altered awareness during seizures^4,32^. While survey-based scales of seizure severity are effective in capturing associated behaviors^12^, they lack a measure of how brain networks are altered by epilepsy. Emerging evidence suggests that structural^30^ and functional^33^ networks are altered in people with severe seizures and epileptic networks acutely generate seizures that manifest as different network dynamics^34^. Our measure for seizure severity, which uncovers differences between seizures within and across individual patients, builds on existing seizure classification and severity scales by quantifying electrographic duration and spread—both characteristics of widespread, pathological brain networks.

Epilepsy duration, response to medication taper, and surgical outcomes are clinical variables that independently measure properties of epilepsy burden. Epilepsy duration associates with widespread abnormalities in the epileptic network^22^ and quality of life^5^, though some patients, including those with genetic seizure disorders, do not experience progressively worse seizures with epilepsy duration. We observe a weak, positive correlation between patient-level seizure severity and epilepsy duration in accordance with previous findings, perhaps limited by the small sample size and heterogeneity of our cohort. Moreover, our finding that more severe seizures occur at lower ASM loads is robust to various taper strategies determined by clinical need. A growing body of evidence suggests that rapid seizure spread and a propensity for FBTCS are guided by a diffuse epileptic network and indicate poor surgical outcome^35,36^. We find that the seizure severity score similarly measures properties of the epileptic network that manifest as poor surgical outcome. Despite substantial heterogeneities in the patient population, our findings that seizure severity is associated with epilepsy duration, response to medication taper, and surgical outcome are promising for determining a clinical biomarker that holistically measures epilepsy burden.

The current treatment approach for severe seizures in the EMU is emergency administration of short-acting benzodiazepines such as lorazepam (Ativan) which acutely reduces or terminates seizures^37^. Severe seizures in this setting are detected by visual observation of specific semiological signs by clinical experts, and there is no indication of severe seizures prior to onset. Previous work has indicated that iEEG network properties may differ for moderate and high severity seizures^31,38^. Our results show that network changes before seizure onset may predict severity within individual patients. However, the patient-specific nature of our result indicates that several mechanisms may cause severe seizures, and not all are detected with the sparse spatial sampling of iEEG within the controlled EMU setting. Seizure triggers from environmental factors, sleep stages, and other behavioral changes which are inadequately captured in iEEG recordings may also affect the severity of an impending seizure. These factors must be integrated with iEEG network measures to build a robust seizure severity predictor. We envision an application of the seizure severity predictor where a patient’s propensity for severe seizures may dictate their medication prescriptions and taper strategy in the EMU, and clinicians and caregivers can intervene with acute medication to curb severe seizures within a warning period to ensure patient safety.

Our work has several important clinical implications. Neurostimulation-based therapies such as deep-brain stimulation (DBS)^39^ and responsive neurostimulation (RNS)^40^ rely on iterative adjustments of stimulation parameters such as amplitude, frequency, and pulse width to effectively reduce seizures. Searching the parameter space of stimulation devices to optimize therapy remains an outstanding challenge^40^. Measuring changes in seizure severity following stimulation parameter adjustments could close the loop for improving iterative therapies and enable clinicians to approach optimal device settings more rapidly. The emerging development of minimally invasive devices with more recording channels and a suite of on-board algorithms could leverage seizure severity recordings to self-optimize parameters without manual clinical input^41–43^. Administration of ASMs are also iterative, as dosage and new medications are altered based on patient reports of seizure frequency^44^. Changes in seizure severity could objectively measure the effects of medication administration and decrease the time to finding effective medication plans for patients. Previous studies have indicated that subclinical seizures, which are not captured in seizure diaries, may affect mood and impair cognition^45^. Detecting and incorporating the severity of subclinical seizures and interictal epileptiform activity in the overall burden of epilepsy could guide therapies to diminish deleterious effects on mood and cognition. Bringing quantitative measures of seizure severity into therapies of neurosurgery, neuromodulation, and medication might better measure one’s epilepsy condition over time.

Though our study outlines a novel methodology for measuring seizure severity, certain limitations must be noted. Intracranial EEG is implanted with the goal of identifying the epileptogenic zone and patients do not have a uniform number of electrodes nor a uniform number of sampled regions. We chose two methodological considerations to mitigate this limitation. First, we measured seizure spread as the number of brain regions where at least one electrode in a counted region was recruited to the seizure to account for spatial oversampling of the hypothesized seizure onset zone. We chose not to normalize the number of recruited regions by the number of implanted regions since some subjects had unilateral implants only. Second, we summarized the preictal data with node strength, a global network measure that is robust to the incomplete sampling of the iEEG brain network, ^46,47^. Baseline seizure frequency and severity were not controlled for when assessing an individual’s propensity for severe seizures following medication taper, and future studies might additionally incorporate an individual’s trajectory in seizure severity over the course of their disease using clinical notes from the electronic health record^48^. People admitted to the EMU are often medically refractory, thus seizure severity scores may differ in ambulatory EEG, where an individual may be exposed to more seizure triggers. We have developed and validated a quantitative seizure severity score using iEEG data as a proof of principle, and future work should extend these concepts to non-invasive EEG and chronic, ambulatory iEEG devices to measure long-term changes in seizure severity. Despite these limitations, our proposed framework for calculating seizure severity can be applied across a multi-center dataset of seizures to overcome center-specific biases in clinical practice.

## Conclusion

We developed and validated a continuous measure for seizure severity that combines seizure duration, spread, and semiology. We show that the seizure severity score detects variability in seizures within and across patients and is sensitive to measures of epilepsy severity. Finally, we propose a preictal iEEG network measure that associates with seizure severity. As the number of palliative measures for drug-resistant epilepsy grows^7,49^, new metrics are critical to evaluate epilepsy burden following medication administration, neuromodulation, and neurosurgery. By proposing a novel, quantitative metric for seizure severity, we seek provide an objective measure for epilepsy burden and improve clinical care for people with epilepsy.

## Data Availability

We have made all our code available at https://github.com/penn-cnt/Pattnaik-seizure-severity. All iEEG recordings are accessible from iEEG.org.

https://www.ieeg.org/

## Acknowledgements

We thank Jacqueline Boccanfuso, Magda Wernovsky, and Anjali Ravikanti for assistance with data organization.

## Author contributions

**Conceptualization:** AP, NS, BL; **Data Curation:** AP, NG, IO, AR, WO, JB, EC, NS**; Formal Analysis:** AP, NG, IO, NS**; Funding Acquisition:** BL**; Investigation:** AP, NS, BL**; Methodology:** AP, NS, BL**; Project Administration:** AP, NS, BL**; Software:** AP, NS, IO, AR, NS**; Supervision:** KD, NS, BL**; Writing – Original Draft Preparation:** AP, NS, BL**; Writing – Review & Editing:** AP, NG, IO, AR, WO, BS, JB, EC, SS, KD, NS, BL

