## Supporting Information for "A quantitative tool for seizure severity: diagnostic and therapeutic applications"

**Non-negative matrix factorization to calculate severity**

Each seizure was represented by the three features, and we performed a data-driven dimensionality reduction using non-negative matrix factorization (NMF). NMF decomposes a feature matrix *A* with dimensions [*n_samples_* x *n_features_*] into two non-negative matrices: a coefficient matrix *W* with dimensions [*n_samples_* x *n_components_*] and a component matrix *H* with dimensions [*n_components_* x *n_features_*] such that $A \approx WH = \hat{A}$. To ensure stable solutions, we initialized NMF with Nonnegative Double Singular Value Decomposition. Zeros in the sparse initialization were filled with the average of $A$. We used a multiplicative update solver to decompose A while minimizing the Frobenius norm of $\hat{A}$. We set *n_components_* to 1, to perform a dimensionality reduction from 3 features to 1 dimension. We interpreted the vector *W* as the severity value of each seizure and *H* as the vector of weights applied to each of the three features. NMF combines seizure duration, spread, and semiology into a scalar that we interpret as seizure severity.

**Absolute slope seizure detector**

Methods are adapted from a previous publication^1^. First, absolute slope $S_{i}\left( t \right)$was computed for each channel *i* as

$$S_{i}\left( t \right)=\left| \frac{\Delta EEG_{i}\left( t \right)}{\Delta t} \right|,$$

where $\Delta EEG_{i}\left( t \right)$ is the difference in voltage between consecutive values$,$ and $\Delta t$ is the sampling timestep. The standard deviation of absolute slope in channel *i* across interictal clips, $\sigma_{i}^{ref},$ was calculated to approximate a patient-specific baseline and was used to normalize the absolute slope of the ictal clip ($\tilde{S_{i}}\left( t \right))$:

$$\tilde{S_{i}}\left( t \right)=S_{i}\left( t \right)/\sigma_{i}^{ref}, with$$

$$\sigma_{i}^{ref}=std\left( S_{i,interictal} \right).$$

Next, the ratio of time samples above a patient-specific absolute-slope threshold ($S_{threshold}$) compared to the total number of time samples in the seizure, T, was computed for each channel *i* as

$$w_{i}=\frac{\sum_{t=1}^{T} 1_{\tilde{S_{i}}\left( t \right)>S_{threshold}}}{T}.$$

Finally, a window threshold ($w_{threshold}$) was applied to separate channels with persistent seizure activity from channels with noise or brief epileptogenic spikes. The vector ***r*** consists of binary labels for each channel as recruited or non-recruited into the seizure, with each element$r_{i}$ calculated as

$$r_{i}=1_{w_{i}>w_{threshold}}.$$

Seizure spread is defined as the number of brain regions sampled by channels where $r_{i}$ = 1.

**Supplementary tables and figures**

**Table S1.** **Extended patient table with clinical variables**

| **Patient** | **1- year Engel outcome** | **Therapy** | **Implant** | **Target** | **Laterality** | **Lesion status** | **Gender** | **Epilepsy duration** |
| --- | --- | --- | --- | --- | --- | --- | --- | --- |
| HUP064 | 1.4 | Resection | ECoG | Frontal | L | Lesional | M | 18 |
| HUP065 | 1.1 | Resection | ECoG | Temporal | R | Lesional | M | 34 |
| HUP068 | 1.1 | Resection | ECoG | Temporal | R | Non-Lesional | F | 15 |
| HUP070 | 1.2 | Resection | ECoG | FP | L | Non-Lesional | M | 21 |
| HUP073 | 1.4 | Resection | ECoG | Frontal | R | Non-Lesional | M | 35 |
| HUP074 | 1.1 | Resection | ECoG | Temporal | L | Lesional | F | 20 |
| HUP075 | 2.4 | Resection | ECoG | Temporal | L | Non-Lesional | F | 5 |
| HUP078 | 2.1 | Resection | ECoG | Temporal | L | Lesional | M | 54 |
| HUP080 | 2.2 | Resection | ECoG | Temporal | L | Non-Lesional | F | 6 |
| HUP083 | 1.1 | Resection | ECoG | Parietal | L | Lesional | M | 21 |
| HUP086 | 2.1 | Resection | ECoG | Temporal | L | Non-Lesional | F | 8 |
| HUP087 | 1.1 | Resection | ECoG | Frontal | L | Lesional | M | 5 |
| HUP088 | 1.4 | Resection | ECoG | Temporal | L | Lesional | F | 34 |
| HUP089 | 1.2 | Resection | ECoG | Temporal | R | Lesional | M | 6 |
| HUP094 | 1.2 | Resection | ECoG | Temporal | R | Non-Lesional | F | 28 |
| HUP105 | 1.1 | Resection | ECoG | Temporal | R | Lesional | M | 12 |
| HUP106 | 1.2 | Resection | ECoG | Temporal | L | Non-Lesional | F | 21 |
| HUP107 | 1.1 | Resection | ECoG | Temporal | R | Non-Lesional | M | 31 |
| HUP111 | 1.2 | Resection | ECoG | Temporal | R | Non-Lesional | F | 12 |
| HUP112 | 3.1 | Ablation | SEEG | Frontal | R | Lesional | F | 21 |
| HUP114 | 3.1 | Ablation | ECoG | MTL | R | Non-Lesional | F | 20 |
| HUP116 | 1.1 | Ablation | SEEG | MTL | R | Lesional | F | 17 |
| HUP123 | 1.1 | Resection | ECoG | Temporal | R | Lesional | M | 3 |
| HUP125 | 1.1 | Ablation | ECoG | Temporal | L | Non-Lesional | M | 9 |
| HUP126 | 1.1 | Ablation | ECoG | MTL | L | Non-Lesional | F | 4 |
| HUP130 | 1.1 | Ablation | SEEG | MFL | L | Non-Lesional | F | 26 |
| HUP132 | 3.1 | Ablation | SEEG | Temporal | L | Non-Lesional | F | 22 |
| HUP133 | 3.1 | Ablation | SEEG | MTL | L | Non-Lesional | F | 5 |
| HUP134 | 1.2 | Resection | SEEG | Frontal | R | Lesional | M | 24 |
| HUP135 | 2.1 | Ablation | SEEG | MTL | R | Non-Lesional | M | 3 |
| HUP138 | 2.1 | Ablation | SEEG | MTL | L | Lesional | M | 9 |
| HUP139 | 1.1 | Ablation | SEEG | Parietal | L | Lesional | M | 20 |
| HUP140 | 1.2 | Ablation | SEEG | MTL | L | Non-Lesional | F | 21 |
| HUP142 | 1.2 | Ablation | SEEG | MTL | L | Lesional | M | 15 |
| HUP144 | 1.4 | Resection | SEEG | Temporal | R | Lesional | M | 26 |
| HUP146 | 1.1 | Resection | SEEG | Temporal | R | Non-Lesional | M | 12 |
| HUP148 | 1.1 | Ablation | SEEG | Temporal | L | Lesional | M | 7 |
| HUP150 | 1.1 | Ablation | SEEG | Insular | R | Lesional | M | 13 |
| HUP151 | 2.1 | Ablation | SEEG | MFL | R | Non-Lesional | M | 27 |
| HUP157 | 1.2 | Ablation | SEEG | MTL | L | Non-Lesional | M | 9 |
| HUP158 | 3.1 | Ablation | SEEG | Insular | R | Non-Lesional | M | 25 |
| HUP162 | 3.1 | Ablation | SEEG | MTL | L | Non-Lesional | F | 14 |
| HUP163 | 1.4 | Ablation | SEEG | MTL | L | Non-Lesional | F | 10 |
| HUP164 | 1.4 | Ablation | SEEG | MTL | L | Lesional | F | 20 |
| HUP166 | 3.1 | Resection | SEEG | Temporal | L | Lesional | M | 22 |
| HUP171 | 2.1 | Ablation | SEEG | Frontal | L | Non-Lesional | M | 44 |
| HUP172 | 1.1 | Ablation | SEEG | Frontal | L | Non-Lesional | F | 26 |
| HUP177 | 1.1 | Resection | SEEG | Temporal | R | Non-Lesional | F | 37 |
| HUP181 | 3.1 | Ablation | SEEG | Temporal | L | Lesional | F | 15 |
| HUP185 | N/A | Ablation | SEEG | MTL | L | Lesional | M | 29 |
| HUP187 | 1.2 | Ablation | SEEG | MTL | R | Non-Lesional | M | 7 |
| HUP188 | 3.1 | Resection | SEEG | Frontal | L | Lesional | F | 15 |
| HUP190 | 3.1 | Resection | SEEG | MTL | L | Non-Lesional | M | 13 |
| HUP191 | N/A | Resection | SEEG | MTL | L | Lesional | F | 16 |

**Table S2.** **National Hospital Seizure Severity Scale**^2^

| How often has the patient fallen to the ground in this type of seizure? | Nearly always or always | 4 |
| --- | --- | --- |
|  | Often | 3 |
|  | Occasionally | 2 |
|  | Never | 0 |
| Has this type of seizure caused any of the following? (score only the worst) | Burns, scalds, deep cuts, fractures | 4 |
|  | Bitten tongue, severe headaches | 3 |
|  | Milder injuries, mild headache | 2 |
|  | No injuries | 0 |
| How often has the patient been incontinent of urine in this type of seizure? | Nearly always or always | 4 |
|  | Often | 3 |
|  | Occasionally | 2 |
|  | Never | 0 |
| If the seizure causes loss of consciousness, is there a warning long enough for the patient to protect him/herself? (no loss of consciousness or seizures only while asleep scores 0) | Never | 2 |
|  | Sometimes | 1 |
|  | Nearly always or always | 0 |
| How long is it until the patient is really back to normal after the seizure? | Less than 1 min | 0 |
|  | Between 1 and 10 min | 1 |
|  | Between 10 min and 1 h | 2 |
|  | Between 1 and 3 h | 3 |
|  | More than 3 h | 4 |
| Do the following events occur in this type of seizure? | Seriously disruptive automatisms (e.g., shouting, wandering, undressing) | 4 |
|  | Mild automatisms or focal jerking | 2 |
|  | None | 0 |
| Add 1 point |  | 1 |

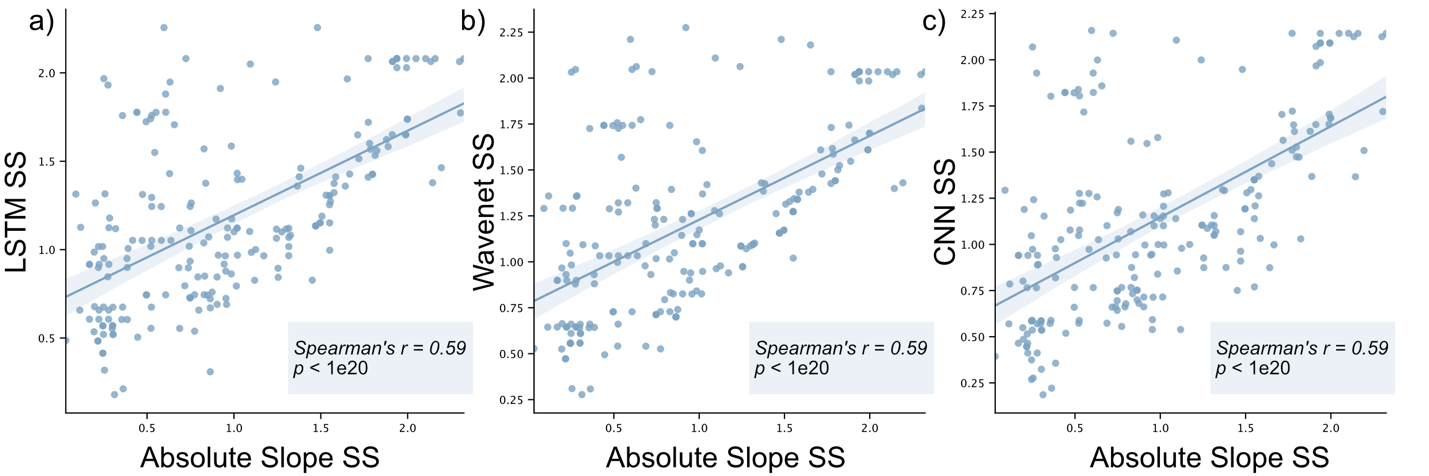

**Figure S1. Seizure severity scores are consistent across seizure spread methods** We replicated our seizure severity methods using three alternative, deep-learning based methods to calculate seizure spread: a) a Long short-term memory (LSTM) architecture, b) a Wavenet architecture, and c) a one-dimensional Convolutional Neural Network (CNN). Models were trained to classify clips as ictal or interictal, and the probability of classification was obtained to estimate each channel’s likelihood for being part of the seizure at each time point. Detailed information about the models can be found elsewhere^3^. We determined thresholds on the seizure probability to classify each channel as seizure or non-seizure and used the same localization pipeline outlined in Methods to determine the number of seizing regions. Seizure severity was re-calculated using each seizure spread model we and compared these new model-based seizure severity scores with the original seizure severity scores reported in the main manuscript.

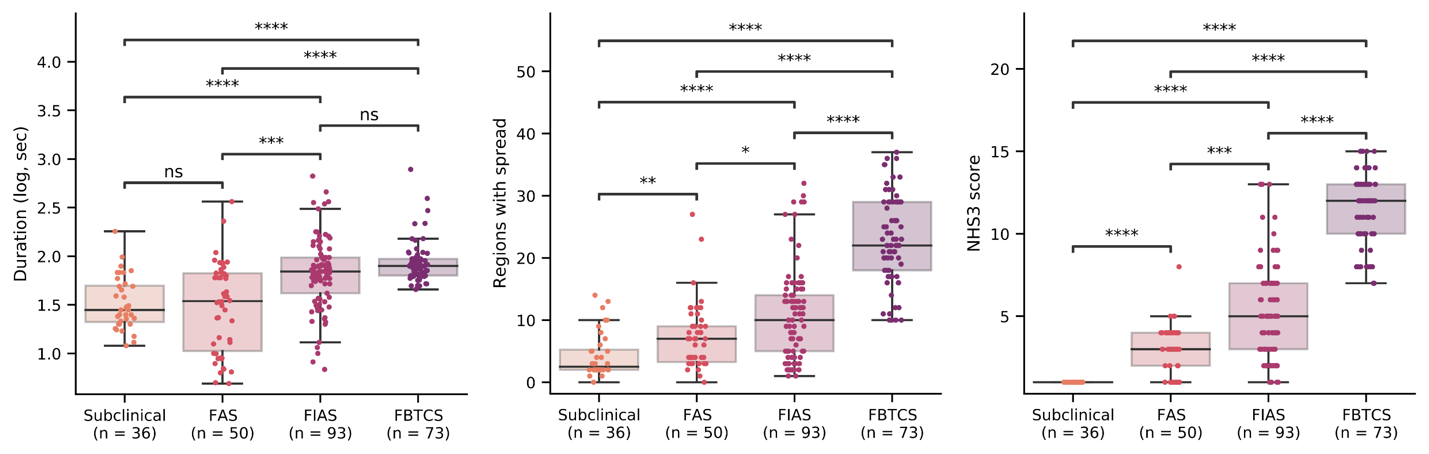

**Figure S2. Components of seizure severity separate across seizure type** We compared each component of the seizure severity score across seizure type to determine if certain features were driving our validation (Kruskal-Wallis test with post-hoc Dunn’s test, *p* <0.05 for all pairs).
